# Validation of Gait Tasks in SynapTrack Mobile App for Cervical Spondylotic Myelopathy

**DOI:** 10.64898/2026.05.27.26354225

**Authors:** Abigail Lewis, Faraz Arkam, Benjamin Steel, Erdong Chen, Pranay Singh, Salim Yakdan, Isaac Becker, Wenwen Guo, Ahmed Shahrabani, Philip RO Payne, Zoher Ghogawala, Michael P. Steinmetz, Brian Neuman, Wilson Z. Ray, Ryan Duncan, Jacob Greenberg

## Abstract

**Background:** Gait impairment is a central sign of cervical spondylotic myelopathy (CSM) that is typically evaluated through subjective patient-reported questionnaires or objective in-clinic measures. These systems require substantial resources to administer and are poorly suited for longitudinal monitoring, however, emerging smartphone applications present an efficient alternative. We developed and assessed the validity of a data processing framework based on the SynapTrack smartphone application to assess gait function in individuals with CSM.

**Methods:** Participants completed walking tasks which were recorded on both the SynapTrack app and a gold standard gait mat. Acceleration data extracted from the smartphone by the app were filtered and processed to produce gait cycle features including velocity, step time, waveform features and frequency domain features. Standard gait features were compared across the two methods by correlation and Bland-Altman plots to assess validity. App-based gait features were then compared to the standard modified Japanese Orthopedic Association (mJOA) assessment to determine construct validity through correlation and ability to discriminate between individuals with CSM and healthy controls. Finally, intraclass correlation coefficients were used to measure test-retest reliability across app features.

**Results:** A total of 110 participants were included in this study, of which 55 (50%) had CSM, 24 (22%) had peripheral neuropathy, and 31 (28%) were healthy controls. SynapTrack gait measures including velocity, step time, and double support showed strong validity as indicated through Bland-Altman plots and high correlation (>0.8) with mat features. In addition to the gait features, acceleration root mean square, acceleration crest, spectral entropy, and dominant frequency showed strong construct validity compared to the mJOA across correlation (0.2-0.54), trend test (p < 0.001), and AUROC (0.62-0.79) analyses. ICCs showed moderate test-retest reliability (0.55-0.71).

**Discussion:** The proposed framework for processing gait data showed strong validity compared to the gold standard mat and high construct validity compared to the mJOA suggesting the utility of the SynapTrack app as an efficient alternative to existing methods. The confirmation of gait metrics related to CSM severity and identification of relevant waveform and frequency domain features present opportunities to use smartphone apps to develop ecologically valid data driven markers of CSM severity.

## Introduction

Cervical spondylotic myelopathy (CSM) refers to chronic, symptomatic spinal cord compression.(1,2) Although prevalence is difficult to quantify and often underestimated,(3) it is estimated that CSM impacts approximately 2% of adults.(4,5) Severe neurological deficits, including impaired dexterity and declines in gait and balance, commonly accompany CSM.(2) However, the broad range of symptoms makes CSM progression challenging to monitor.

Assessment of CSM progression typically relies on in-clinic testing and patient reported outcomes through clinically delivered questionnaires which occur with a clinical visit cadence of every three to twelve months.(1,2) The conventional clinically administered questionnaire used to monitor CSM progression is the modified Japanese Orthopedic Association Score (mJOA).(6– 11) The subjective nature of questionnaire responses and infrequent administration limit their utility in objectively assessing and identifying subtle, short-term changes in CSM severity. Thus, objective measures of CSM severity related to neurological abilities have shown promise in improving detection of CSM progression. However, objective measures also remain challenging to collect and scale due to substantial resource requirements and, typically, similar reliance on in-clinic visits.

Gait impairment is one of the most common signs of CSM and its correlation with disease severity has been consistently validated in the literature.(9,12–17) As such, gait assessment potentially presents a promising approach to measure CSM severity. While quantified subjectively in CSM questionnaires, gait has also been objectively measured through multiple approaches, including laboratory-based video analysis, floor sensors, and wearable devices like inertial measurement units (IMU) or active markers.(18–23) These systems, though, are poorly suited for longitudinal monitoring, require substantial resources to administer, and require patient travel to centralized testing locations. Better tools are needed to measure gait function that are both efficient and objective.

More recently, mobile health approaches relying on embedded smartphone IMUs have been identified as a valid and reliable mechanism to objectively assess physical functioning, and gait specifically, while addressing the resource and temporal constraints of in-office assessment.(18,24–28) The flexibility of smartphone-based disease severity measurements supports remote patient monitoring in real-world circumstances at a frequency from which subtle changes in gait ability can be detected. Although apps are being developed for general gait analysis and in the contexts of Parkinson’s Disease and multiple sclerosis, there is limited literature validating the core analytic approaches used to extract gait metrics in these diseases, with scarce efforts applied to CSM specifically.(29–31) Similarly, existing apps for CSM assessment have undergone minimal analytical validation and have only been piloted in small sample sizes (e.g., 7-27 individuals).(32,33) More broadly, sample sizes for validating app-based gait assessments have generally been small and typically restricted to healthy participants. Likewise, data processing details have been sparse, limiting reproducibility and the potential for external validation.(18) These shortcomings present major barriers to scaling reliable, trustworthy digital gait monitoring using widely available consumer smartphone technology.

To address this broader methodologic gap and the specific shortcomings of CSM monitoring, our team recently developed a smartphone application, SynapTrack, to assess neurological abilities in individuals with CSM. To provide a foundation for this work and to guide the broader field of digital gait monitoring, this study assesses the performance of a processing framework for extracting gait characteristics from the SynapTrack app. Through assessing the validity, reliability, and construct validity of gait metrics, we provide evidence for their use in CSM and a guide to smartphone-based gait analysis of neurological function.

## Methods

This study was conducted from March 2025 through March 2026 and was approved by the Washington University School of Medicine (WUSM) IRB (202401026).

Participants with CSM were recruited through elective clinical practices at a tertiary medical center, and controls were recruited through public advertising and research volunteer services. We additionally recruited a small number of participants with peripheral neuropathy to explore the generalizability of SynapTrack measures across clinical populations. Following enrollment, participants completed an in-person assessment in which they performed a standard assessment of gait function in neurologic care, in addition to SynapTrack based assessments. During the gait task, participants were instructed to walk in a straight line for 12 steps similar to other app-based gait task instructions approximating the standard 25-Foot Walk.(14,28,34,35) Motion during the task was simultaneously recorded by the SynapTrack app and the ProtoKinetics Zeno Movement (PKM) mat which was subsequently used as a gold standard comparison. After the initial in-person testing, some participants were instructed to repeat the gait task at home, typically up to 4 days per month, though varying depending on clinical care pathways (e.g., operative vs. nonoperative patients). The full workflow can be seen in **Figure 1**.

**Figure 1.**
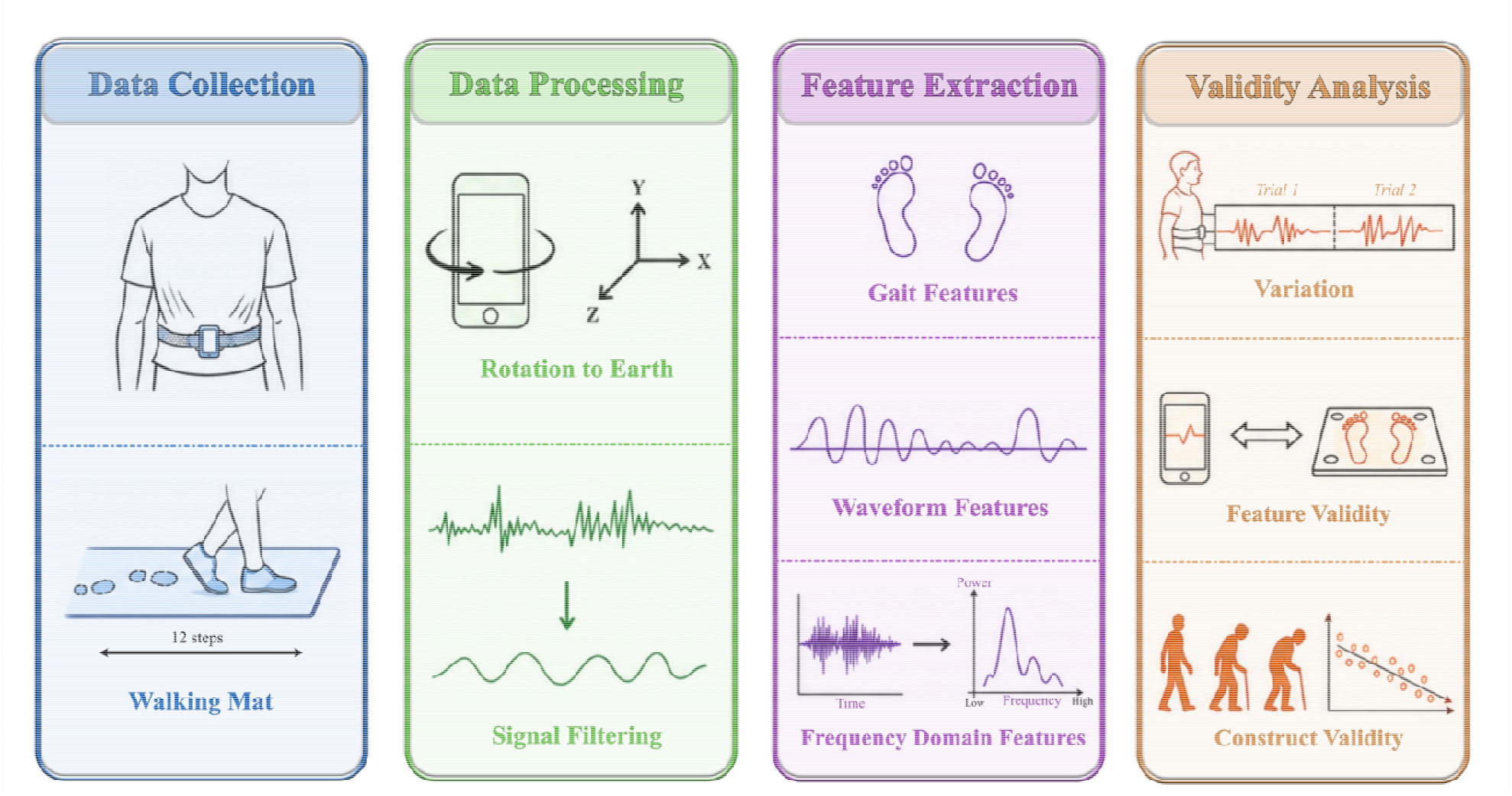
Workflow diagram.

### Smartphone Data Processing

IMU data for each gait task were collected by the SynapTrack app using the built-in smartphone IMU sensors. Data included tri-axial acceleration, tri-axial gyroscope, and quaternion measures recorded at 100 Hz for the duration of the task. The acceleration signals were adjusted to remove gravitational acceleration and projected into Earth’s coordinates using the quaternion resulting in two sets of triaxial acceleration signals (phone-oriented and Earth oriented). To address noise in the acceleration signals imposed by the sensors, acceleration signals were filtered with a fourth order low-pass Butterworth filter to remove noise in the data unrelated to participant movement. In gait analysis, typical threshold values for this filter range from 2 to 20 Hz. Thus, we tested thresholds between 2 and 20 Hz with a 2 Hz increment and identified the optimal threshold by the maximized validity of extracted gait features.

Walking bouts were extracted for each task to remove excess standing time during initiation and end of the task. Methods from Straczkiewicz, Huang, and Onnela were used to extract walking bouts in which absolute acceleration thresholding is used to determine when walking is initiated.(36) Direction of forward movement was defined as the first principal component of the acceleration signals in the X and Y directions according to Earth’s orientation.(37)

### Gait Cycle Features

Features of the gait cycle can be used to assess gait functionality and are often defined using specific components of the gait cycle. The gait cycle is typically defined by heel strikes (HS), which refer to initial contact of a heel with the ground, and toe offs (TO), which refer to final contact of a toe with the ground. A stride is defined by the time between HS of the same foot while a step is defined by the time between HS of opposite feet. Single and double support time are respectively defined by having one or two feet connected with the ground. Single support time is calculated as the time between the TO and HS of the same foot, while double support time is calculated as the time between the HS of one foot and the subsequent TO of the opposite foot. HS and TO were approximated by maxima and minima in the acceleration signal in the anterior-posterior axis.(18,38)

Extracted features and the methods of calculation are listed in **Table 1**. Gait cycle features were categorized as traditional gait features, waveform features, and frequency domain features. Traditional gait features included the standardly assessed step time, double support time, single support time, and velocity which have been previously validated to be associated with CSM severity.(15,18,19,25) Waveform features reflected the structure of the acceleration signals rather than explicit gait constructs. Frequency domain features reflected the wave structure in the frequency domain. Conceptual interpretations of the waveform and frequency features are included in **Supplemental Table 1**.

**Table 1.**
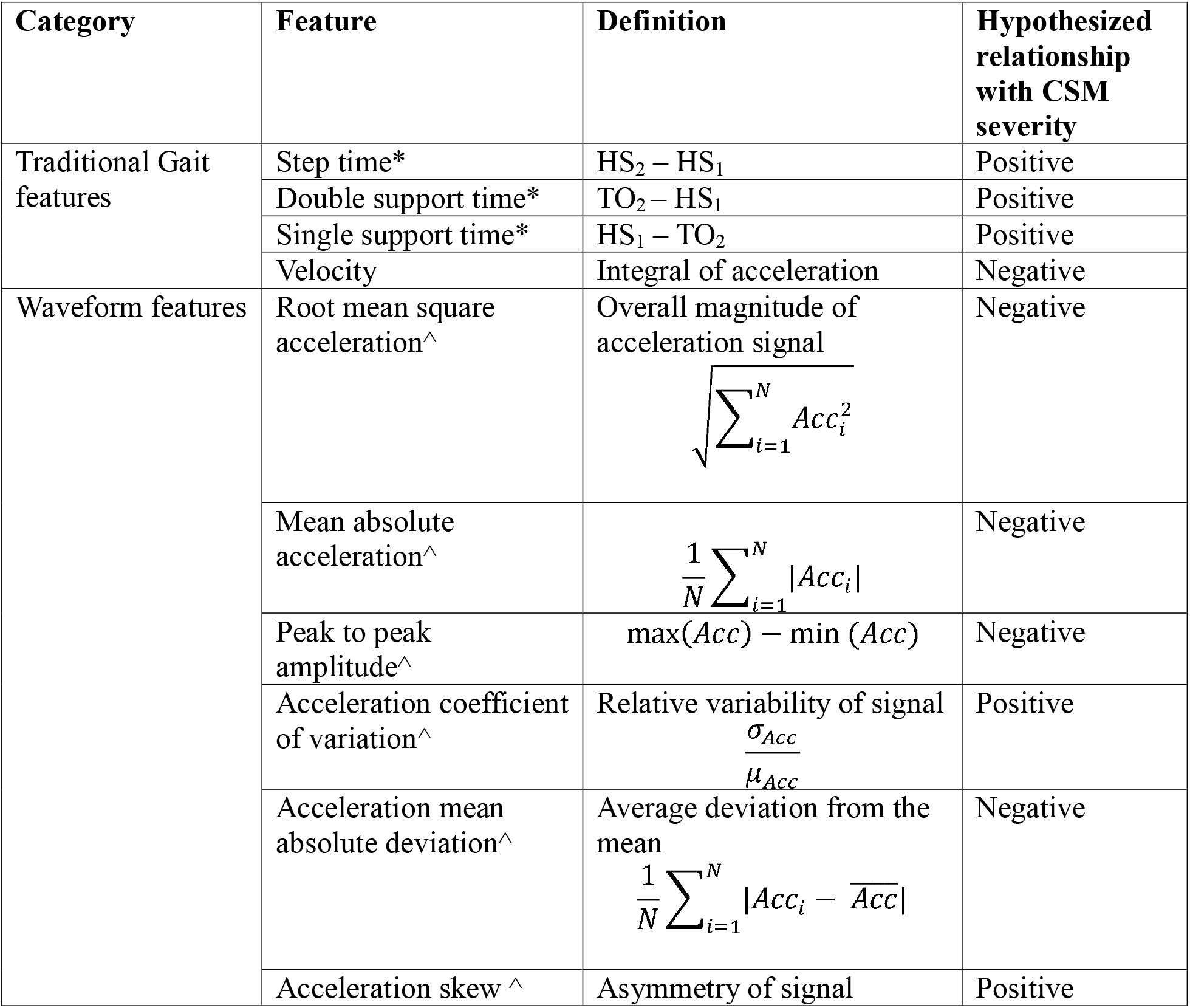
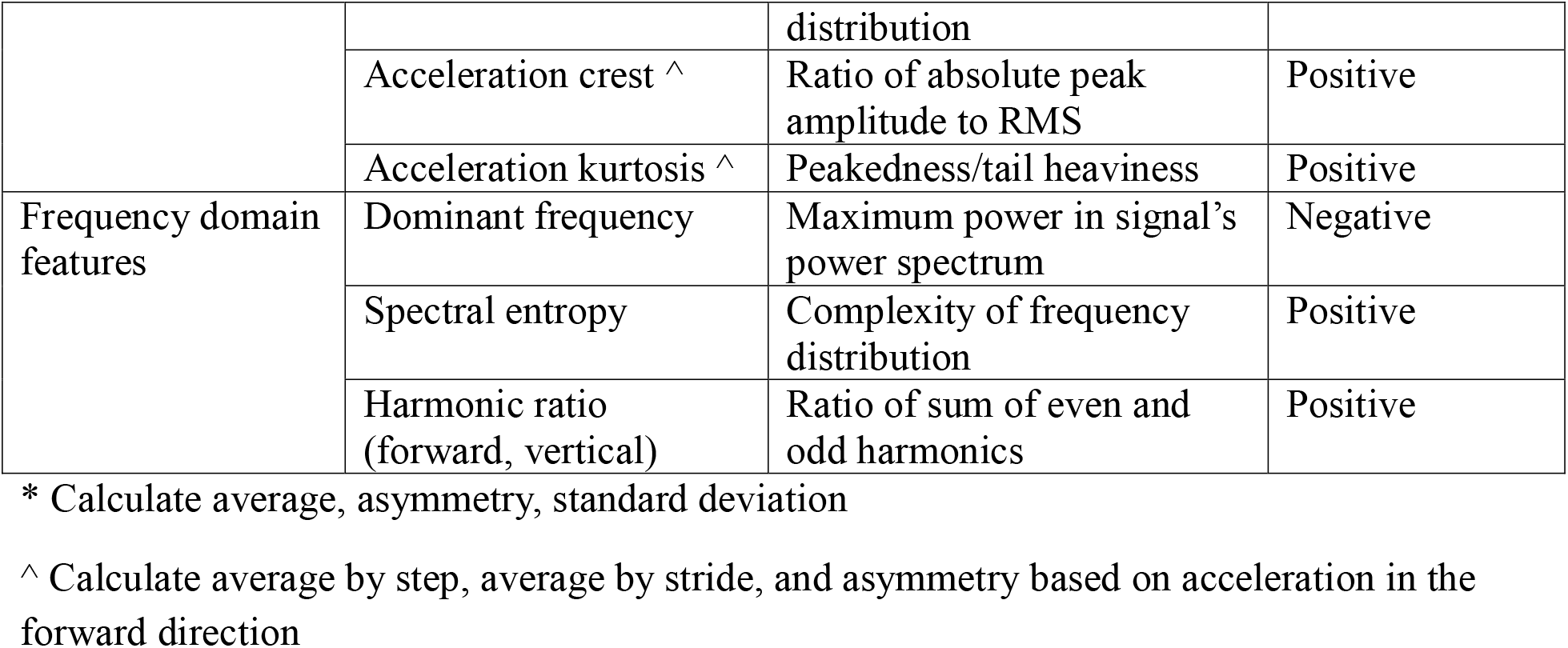
Gait Cycle Features.

### Feature Validity

Validity was assessed for each of the features in the gait category (**Table 1**) by comparison with the gold standard measures recorded by the PKM mat, based on all trials that were completed both on the SynapTrack app and in-person on the PKM mat. Correlation coefficients were calculated comparing the SynapTrack app and PKM mat estimates where correlation greater than 0.8 indicated high validity and correlation between 0.5 and 0.8 indicated moderate validity. Bland-Altman plots were used to additionally assess distributional alignment.

### Construct Validity

Construct validity was determined by comparing gait features extracted from the SynapTrack app to the mJOA scale and assessed through three approaches. The mJOA scale was selected due to its use as the gold standard assessment of CSM severity in clinical care.(7,8,10) First, correlation coefficients were calculated comparing all features listed in **Table 1** to the patient reported mJOA. Second, ordinal trends in distributions of each feature across participant groups with varying CSM severity were tested with Jonckheere-Terpstra (JT) tests. In this case, mild myelopathy was defined by an mJOA score between 15 and 17, moderate myelopathy was defined by an mJOA score between 12 and 14, and severe myelopathy was defined by an mJOA score of 11 or less.(8) The correlation and trend tests were conducted only in participants with CSM, and significant p-values were identified using the Benjamini-Hochberg procedure for false discovery rate (FDR) corrections with an initial cutoff of α = 0.05. Third, areas under the receiver operating characteristic curves (AUROC) were calculated to determine each feature’s ability to discriminate between healthy controls and participants with CSM. Construct validity was identified by a combination of the correlation coefficients, JT trend tests of feature distributions across categories of CSM severity, and ability to discriminate between healthy controls and participants with CSM measured by AUROC.

### Intra-subject Variation

Reliability was first evaluated among participants with at least two app-based gait assessments using a two-way mixed-effects consistency ICC (ICC [3,1]). A secondary analysis was performed among participants with four assessments obtained within a 30-day period. In this analysis, the first two assessments and the subsequent two assessments were averaged to create two repeated measurements per participant, and ICC (3,1) values were calculated using these paired averaged measurements. This approach was used to evaluate whether averaging repeated gait assessments improved measurement reliability. For gait mat features, ICC (3,1) values were calculated using the first two available gait mat assessments obtained within a 30-day period.

## Results

Cohort characteristics including demographics and disease status are shown in **Table 2**. A total of 110 participants were included in this study. Of the 110 participants, 31 (28%) were healthy controls, 55 (50%) had CSM, and 24 (22%) had peripheral neuropathy. Most of the participants were non-Hispanic White, had at least a college degree, and were either working or retired. There was a similar proportion of male and female participants. The average age of healthy controls was slightly lower than that of participants with CSM or other neuropathy, but demographics did not statistically differ between the healthy controls and participants with CSM (**Table 2**).

**Table 2.** Cohort characteristics. mJOA ranges: Mild (15-17), Moderate (12-14), Severe (≤11).

| Participant Characteristic | Participant Type |  |  |  |  |
| --- | --- | --- | --- | --- | --- |
|  | Overall | Control | Myelopathy | Peripheral Neuropathy | P-Value |
| **n** | 110 | 31 (28%) | 55 (50%) | 24 (22%) |  |
| **Race, n (%)** |  |  |  |  |  |
| Missing | 6 (5.5) | 0 (0.0) | 4 (7.3) | 2 (8.3) | 0.191 |
| White | 92 (83.6) | 27 (87.1) | 44 (80.0) | 21 (87.5) |  |
| Black | 6 (5.5) | 0 (0.0) | 5 (9.1) | 1 (4.2) |  |
| Asian | 5 (4.5) | 3 (9.7) | 2 (3.6) | 0 (0.0) |  |
| Native American | 1 (0.9) | 1 (3.2) | 0 (0.0) | 0 (0.0) |  |
| **Ethnicity, n (%)** |  |  |  |  |  |
| Missing | 8 (7.3) | 1 (3.2) | 5 (9.1) | 2 (8.3) | 0.494 |
| Not Hispanic or Latino | 3 (2.7) | 2 (6.5) | 1 (1.8) | 0 (0.0) |  |
| Hispanic or Latino | 99 (90.0) | 28 (90.3) | 49 (89.1) | 22 (91.7) |  |
| **Gender, n (%)** |  |  |  |  |  |
| Missing | 8 (7.3) | 1 (3.2) | 4 (7.3) | 3 (12.5) | 0.571 |
| Male | 49 (44.5) | 15 (48.4) | 22 (40.0) | 12 (50.0) |  |
| Female | 53 (48.2) | 15 (48.4) | 29 (52.7) | 9 (37.5) |  |
| **Age, mean (SD)** | 59.5 (12.8) | 55.7 (13.3) | 60.0 (13.2) | 63.6 (9.7) | 0.075 |
| **Education Level, n (%)** |  |  |  |  |  |
| Missing | 9 (8.2) | 0 (0.0) | 6 (10.9) | 3 (12.5) | 0.016 |
| Less than high school | 2 (1.8) | 0 (0.0) | 1 (1.8) | 1 (4.2) |  |
| High school degree | 19 (17.3) | 2 (6.5) | 13 (23.6) | 4 (16.7) |  |
| College degree | 46 (41.8) | 13 (41.9) | 19 (34.5) | 14 (58.3) |  |
| Graduate or professional school degree | 34 (30.9) | 16 (51.6) | 16 (29.1) | 2 (8.3) |  |
| Employment Status, n (%) |  |  |  |  |  |
| Missing | 6 (5.5) | 0 (0.0) | 4 (7.3) | 2 (8.3) | 0.080 |
| Actively working | 51 (46.4) | 20 (64.5) | 20 (36.4) | 11 (45.8) |  |
| Homemaker | 2 (1.8) | 1 (3.2) | 0 (0.0) | 1 (4.2) |  |
| Retired | 42 (38.2) | 10 (32.3) | 23 (41.8) | 9 (37.5) |  |
| On Disability | 9 (8.8) | 0 (0.0) | 8 (15.4) | 1 (7.7) |  |
| Disease Severity (mJOA), n (%) |  |  |  |  |  |
| Control | 31 (28.2) | 31 (100.0) |  |  |  |
| Peripheral Neuropathy | 24 (21.8) |  |  | 24 (100.0) |  |
| Mild | 14 (12.7) |  | 14 (25.5) |  |  |
| Moderate | 19 (17.3) |  | 19 (34.5) |  |  |
| Severe | 8 (7.3) |  | 8 (14.5) |  |  |
| Missing | 14 (12.7) |  | 14 (25.5) |  |  |

### Validity

Validity was tested using 141 matched trials for 83 unique participants. A cutoff of 2Hz was selected for filtering as it resulted in the strongest correlations between app measures and gold-standard mat measures (**Table 3**). Using a 2Hz cutoff, app measurements demonstrated strong correlations with the gold-standard mat measurements: velocity (0.84), step time (0.86), single support time (0.56), and double support time (0.82). These correlations combined with the overall trends in the left column of **Figure 2** demonstrate strong validity of app-based measures. Bland-Altman plots (**Figure 2**, right column) further support the validity of the app gait assessments through demonstrated agreement between app and mat measures.

**Table 3.** App validity across filtering thresholds. Table entries show correlations between app and mat measures. (N = 141 trials, 83 individuals)

| Threshold (Hz) | Feature Correlation |  |  |  |
| --- | --- | --- | --- | --- |
|  | Velocity | Step Time | Single Support Time | Double Support Time |
| 2 | **0.84 \*\*\*** | **0.86 \*\*\*** | **0.56 \*\*\*** | **0.82 \*\*\*** |
| 4 | 0.84 *** | 0.82 *** | 0.56 *** | 0.66 *** |
| 6 | 0.84 *** | 0.81 *** | 0.49 *** | 0.61 *** |
| 8 | 0.80 *** | 0.78 *** | 0.47 *** | 0.59 *** |
| 10 | 0.77 *** | 0.76 *** | 0.44 *** | 0.55 *** |
| 12 | 0.77 *** | 0.77 *** | 0.46 *** | 0.57 *** |
| 14 | 0.76 *** | 0.77 *** | 0.45 *** | 0.58 *** |
| 16 | 0.76 *** | 0.75 *** | 0.45 *** | 0.57 *** |
| 18 | 0.76 *** | 0.73 *** | 0.47 *** | 0.55 *** |
| 20 | 0.76 *** | 0.76 *** | 0.49 *** | 0.57 *** |

**Figure 2.**
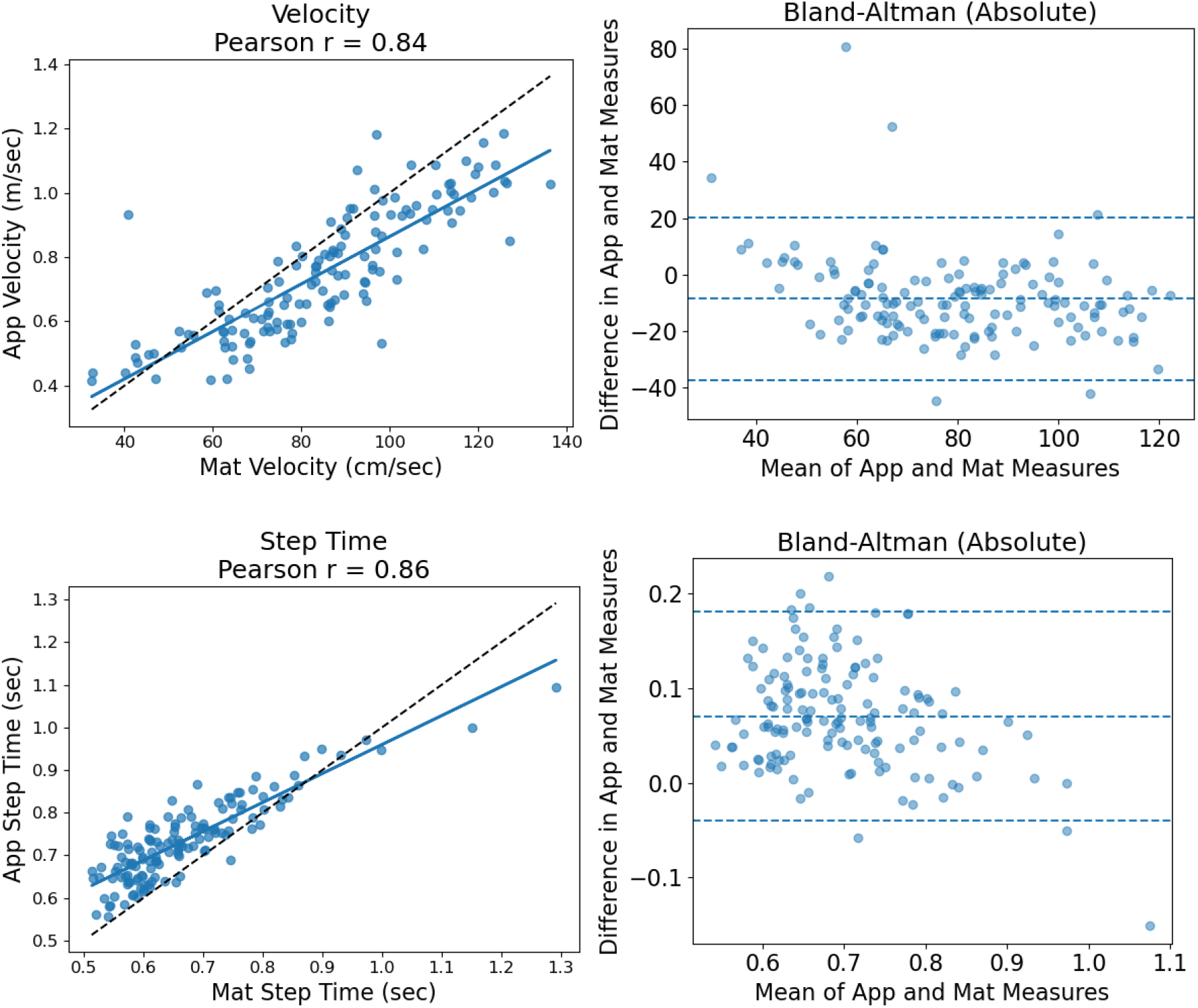
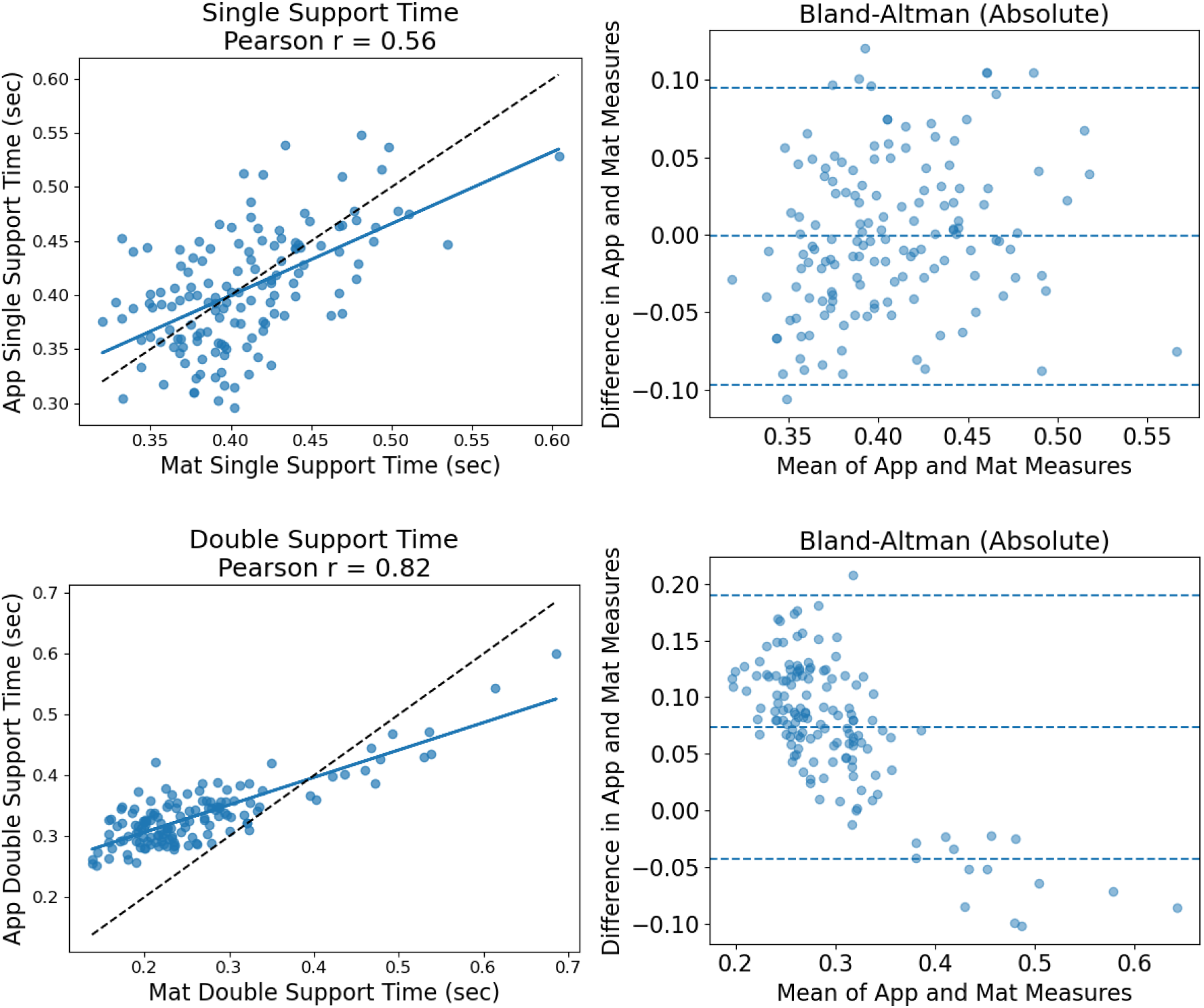
App measure validity. Left column: scatter plots with reference lines. Right column: Bland-Altman plots of absolute agreement. Row order: Velocity, step time, single support time, double support time.

### Construct Validity

A total of 39 features were explored for construct validity. Results for the gait features, waveform features measured at the step level, and frequency features with the strongest construct validity are included in **Table 4**, while results for additional features are included in **Supplemental Table 2**.

**Table 4.** Correlation between gait cycle features and mJOA. Significance is based on FDR adjusted p-values. Bolded features indicate statistically significant agreement with initial hypotheses in Table 1 in at least one of the correlation or trend tests. Correlation and trend tests only include participants with CSM. AUROC calculations include participants with CSM and healthy controls.

| Feature category | Feature | Correlation | Jonckheere-Terpstra Trend Test | AUROC |
| --- | --- | --- | --- | --- |
| **Gait features** | **Step Time** | -0.37*** | 6.12*** | 0.62 |
|  | **Double Support Time** | -0.34*** | 4.79*** | 0.57 |
|  | **Single Support Time** | -0.34*** | 6.46*** | 0.64 |
|  | **Velocity** | 0.41*** | -5.08*** | 0.74 |
| **Waveform features** | **Root Mean Square** | 0.19* | -7.45*** | 0.79 |
|  | **Peak to Peak Amplitude** | 0.19* | -7.38*** | 0.54 |
|  | **Mean Absolute Acceleration** | 0.20* | -7.50*** | 0.65 |
|  | Coefficient of Variation | 0.02 | -1.85 . | 0.57 |
|  | **Mean Absolute Deviation** | 0.20* | -7.61*** | 0.62 |
|  | Skew | -0.07 | -1.73 | 0.60 |
|  | **Kurtosis** | -0.21* | 3.87*** | 0.61 |
|  | **Crest** | -0.32* | 5.56*** | 0.69 |
| **Frequency domain features** | **Dominant Frequency** | 0.31*** | -5.52*** | 0.62 |
|  | **Spectral Entropy** | -0.54*** | 8.23*** | 0.69 |
|  | **Forward Harmonic Ratio** | -0.11 | 2.60* | 0.52 |

Significance Level: . 0.1, \* 0.05, \*\* 0.01, \*\*\* 0.001

All features other than acceleration CV and skew showed a significant relationship with mJOA according to correlation or trend analyses after p-value corrections (**Table 4, Supplemental Table 2**) and matched the hypothesized relationships reported in **Table 1** in participants with CSM.

Absolute correlations between each of the gait features and mJOA were above 0.3 with velocity showing the strongest correlation (0.41) followed by step time (-0.37). Within waveform features, absolute correlations with mJOA ranged from close to zero to 0.32 with crest showing the strongest correlation at -0.32. Within frequency domain features, absolute correlations with mJOA ranged from 0.11 to 0.54 with spectral entropy demonstrating the strongest correlation at - 0.54.

To improve visualization of how gait features varied by known disease groups, we also plotted key gait features stratified by control versus mild, moderate, and severe CSM (**Figure 3**). Results for testing same trend for statistical significance are shown in **Table 4**. Velocity, dominant frequency and RMS were inversely related with myelopathy severity; myelopathy severity decreased with increases in those measures. Double support time, step time, single support time, spectral entropy, and crest were positively correlated with myelopathy severity.

**Figure 3.**
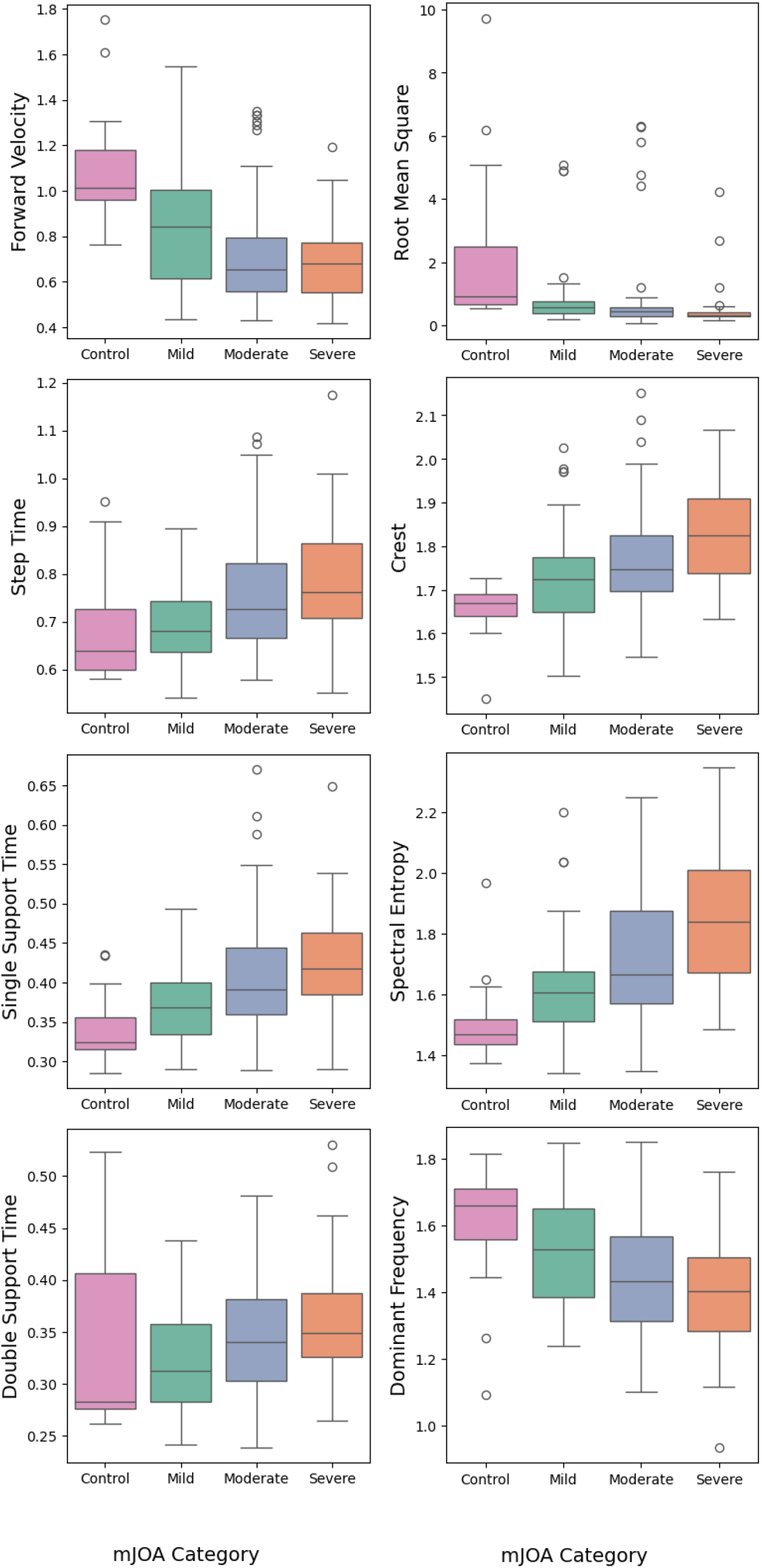
Boxplots of construct validity. Y-axis shows feature value and x-axis shows category of CSM severity based on mJOA score with ranges: Mild (15-17), Moderate (12-14), Severe (≤11).

Many app measures also showed strong capacity for discriminating between healthy controls and participants with CSM (**Figure 4**). AUROC values of the strongest predictors approached 0.8, with many additional features ranging from 0.55 to 0.65. RMS showed the largest discriminative ability with an AUROC of 0.79 followed by forward velocity (AUROC = 0.74), and spectral entropy (AUROC = 0.69). Of note, features from all three categories (gait, waveform, frequency domain) showed promising discriminative ability.

**Figure 4.**
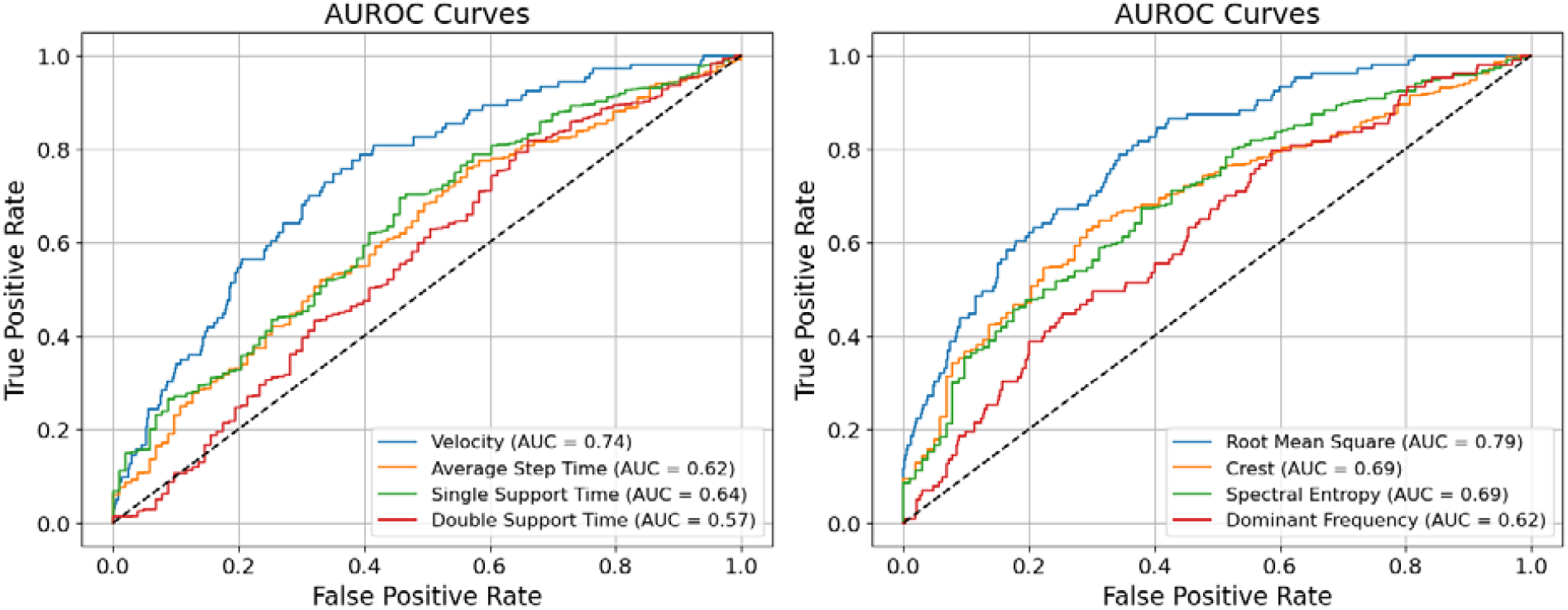
ROC curves using app derived features to identify participants with CSM vs. healthy controls. Panel 1: gait features. Panel 2: proposed features of interest.

Among the waveform and frequency domain features assessed, crest, RMS, spectral entropy, and dominant frequency stood out as promising candidate measures of gait ability across the three measures of construct validity. These four measures showed the highest correlations with mJOA and AUROCs supporting their potential utility as markers in clinical gait assessment.

### Test-Retest Reliability

When using individual gait measurements, ICCs of the gait features ranged from 0.40 (single support) to 0.61 (velocity). However, when averaging two gait trials, ICCs increased, with substantial increases for some measures (e.g. ICC 0.40**→**0.59 for single support) and others having a more moderate change (e.g., ICC 0.61**→**0.71 for velocity), as shown in Table 5. These results indicated that there is natural variability in free-living gait assessments, with repeated gait assessments improving the reliability of this remote testing.

**Table 5:**
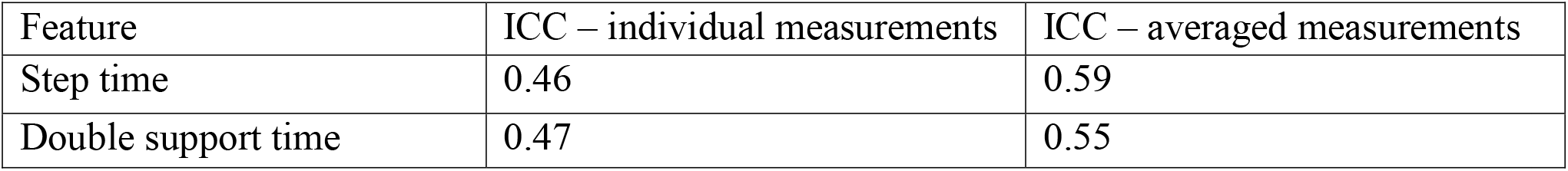
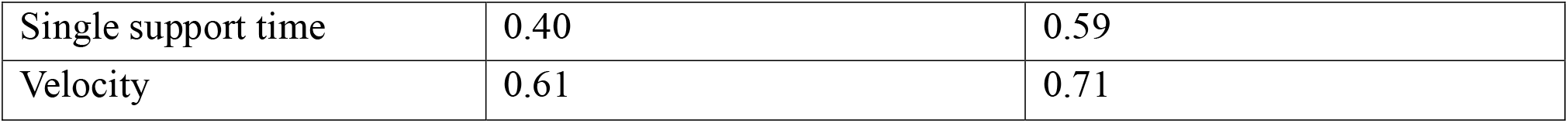
Test-retest reliability of remote gait testing using SynapTrack.

| Feature | ICC – individual measurements | ICC – averaged measurements |
| --- | --- | --- |
| Step time | 0.46 | 0.59 |
| Double support time | 0.47 | 0.55 |
| Single support time | 0.40 | 0.59 |
| Velocity | 0.61 | 0.71 |

## Discussion

In this study we designed a framework to assess gait ability through a walking task on a mobile smartphone app relying on embedded smartphone IMUs. Extracted gait features were compared to measures obtained from the gold standard PKM mat to assess accuracy and features were compared to the mJOA clinical scale to demonstrate construct validity in both healthy controls and participants with CSM. We found that several well-described gait features showed strong analytical validity compared to a gold-standard gait mat, along with construct validity compared to patient-reported CSM. We additionally found strong construct validity across waveform and frequency domain features, suggesting novel, hypothesis-generating metrics reflecting gait quality. These results substantiate the role and methods for remote digital monitoring of CSM and other neurological diseases.

Our gait features demonstrated strong validity compared to the gold standard PKM mat. This supports smartphone evaluation of gait assessment in populations with CSM as a low-cost, efficient alternative to time-gated in-clinic assessments on gait mats that may be cost prohibitive in many contexts. The extracted features were valid across healthy controls and participants with both CSM and peripheral neuropathy in large sample sizes, supporting the generalizability of the results. Double support time showed patterns in the Bland-Altman plot suggesting that errors in our approximation methods were inversely related to actual double support (DS) time; that is, we overestimated DS time for short DS times. However, our approach was more highly correlated than that reported by Apple, and similar agreement patterns have been reported in other studies,(39,40) suggesting potential limits of smartphone-based assessments.

The construct validity assessments demonstrated utility of app derived gait cycle features for measuring CSM severity. These tasks and metrics could be used to monitor disease severity and progression without requiring in-clinic assessment, thereby supporting close disease monitoring while also reducing patient and provider time burdens.(41) Features extracted during these tasks could additionally support development of new clinical scales and biomarkers based on real-world mobility and precise assessments rather than restricted to in-clinic testing and patient-reported severity.

The features from the gait category reflected known relationships between gait cycle function and CSM severity, validating SynapTrack against published literature.(12–14,17,20) In addition, certain waveform and frequency features demonstrated strong construct validity, beyond typical gait features. Although not traditionally defined gait features, many of the waveform and frequency domain metrics encompass aspects of gait related to stability, strength, and balance which are all directly implicated in CSM.

We specifically identified crest, RMS, spectral entropy, and dominant frequency as potentially informative gait features. RMS has been previously explored showing an inverse relationship with general gait ability and CSM severity specifically.(19,23) Based on our review of existing work, crest has not yet been explored in the context of CSM or other neurological diseases. As crest measures extremity of maximum values, it suggests more impulsive or less smooth gait function. Clinically, the positive relationship between crest and CSM severity matches known impairments in CSM gait function. Surprisingly, spectral entropy and dominant frequency showed stronger construct validity than harmonic ratio, despite its validation as an indicator of gait ability in other applications.(19,23) Conceptually, spectral entropy is correlated with walking instability; as the entropy in the frequency domain increases, gait is employing a larger range of frequencies. Thus, the relationship seen between increased spectral entropy and increased CSM severity makes clinical sense. Conversely, dominant frequency measures the maximum power in the frequency signal’s power spectrum which is correlated with velocity. Thus, it expectedly shows a strong relationship with CSM severity. Future work should further validate these measures and explore their clinical utility as indicators of gait function. More broadly, the high construct validity of these novel gait features extracted from smartphone IMU-based data support the utility of this approach in objectively characterizing gait abilities relevant to CSM.

In contrast to assessments of some other apps, we identified moderate within-person variability in gait features in free-living assessment, with ICCs lower than typically reported in similar studies.(42) However, variation in gait is expected during short bouts of activity like the task prescribed in the SynapTrack app, (29,43) particularly in free-living contexts. Further, similar studies have often measured inter-method reliability rather than test-retest reliability. Perhaps most importantly, most other studies have been conducted exclusively in structured laboratory settings.(18) Although initial testing for this study was conducted in a laboratory setting, participants were not instructed to walk at a specific speed and all follow-up testing was completed at home. Thus, our study likely reflects real world variation in gait, especially during short bouts of activities,(29,43) and evaluates natural walking performance. These aspects of our assessments support both ecological and external validity of the SynapTrack gait assessments which may be missing in other published results.

However, gait measures averaged across two observations increased ICCs by a notable degree (e.g., 0.1-0.2), showing how the use of several gait trials can attenuate the influence of real-world gait variability. Due to constraints in our available data, only two observations were used in ICC calculations. However, based on the observed improvements moving from one to two trials, we posit that averaging across three observations, and/or conducting two gait bouts per testing epoch, would further improve test-retest reliability without creating undue patient burden.

The moderate test-retest reliability identified after averaging across observations also raises questions regarding the generalizability of previously reported ICCs near 1 – indicating near perfect reliability – in free-living contexts.(28,30,40) While much of these differences may reflect laboratory versus remote testing, standardized reporting of reliability metrics is also needed, including methods such as outlier handling and window selection, which may impact results. Nonetheless, our encouraging results across healthy controls and participants with CSM and peripheral neuropathy support the extension of this app to disease monitoring in other neurological disorders, like Parkinson’s disease or multiple sclerosis.

### Strengths and Limitations

Our app has been tested in a larger sample size than many published apps, including a balance of healthy controls and participants with CSM and peripheral neuropathy. However, continuing observations in more participants would further validate the clinical utility of this platform. Our sample size in patients with other forms of neuropathy was small, so conclusions on performance specifically in those populations should be tempered. In addition, we observed moderate ranges of intra-participant gait variation. As previously stated, in contrast to other similar studies, we believe this presents a more realistic representation of natural gait variation. However, adjusting the gait task instructions (e.g., “walk as fast as you comfortably can”) may promote higher levels of consistency across trials. Finally, although showing strong validity and construct validity, measures like single support time could benefit from improved extraction methods, and other standard gait measures warrant future investigation (step length, asymmetry measures, etc.).

## Data Availability

Due to concerns related to participant privacy, full data are not available, but reasonable requests for data access will be accommodated when possible.

